# Resurgence of *Bordetella pertussis* in the Netherlands in 2023-2024 reveals novel mutation in T3SS translocator gene *bopD*

**DOI:** 10.1101/2025.09.05.25335159

**Authors:** Linda J. Visser, Janieke van Veldhuizen, Valentijn Tromp, Ramón Noomen, Airien Bonačić-Harpal, Rob Mariman

## Abstract

**Background:** Despite decades of vaccination whooping cough has not been eliminated and epidemics are known to occur every 3 to 5 years. As a consequence of the COVID-19 pandemic and associated non-pharmaceutical interventions, pertussis has been absent in 2020-2022. Many European countries have since seen large pertussis epidemics in 2023 and 2024.

**Aim:** To type Dutch pertussis isolates from 2023-2024 and compare these to pertussis isolates from 2015-2020.

**Methods:** Pertussis isolates received by the reference laboratory are routinely subjected to Illumina sequencing and genotyped. Expression of vaccine antigens and virulence factors was monitored over time (2015-2024). The 2023-2024 pertussis isolates were additionally typed using a cgMLST approach.

**Results:** Between 2015-2020, 98% of isolates belonged to two dominant genotypes. Only 82% of 2023-2024 isolates belonged to these two genotypes, a third genotype was found to make up 13% of recent pertussis isolates. Fewer pertactin-deficient isolates were observed in 2023-2024, compared to isolates from 2015-2020. Furthermore, macrolide resistant isolates have not been detected, in contrast to reports by other European countries. Twenty-one recent isolates were found to contain a novel non-synonymous mutation in the type III secretion system translocator gene, *bopD*.

**Conclusion:** Pertussis resurgence in the Netherlands coincided with the introduction of a third genotype, thereby resulting in increased genetic diversity when compared to isolates from 2015-2020. The third genotype is of the ancestral ptxP1 promoter type. A novel allele of the *bopD* gene has been identified, the implications of this mutation remain unclear at this time.

## Introduction

Whooping cough is an infectious disease caused by *Bordetella pertussis* or *Bordetella parapertussis*. The disease is best known for the characteristic cough, which can last for multiple weeks. Especially for infants the disease can be severe, requiring hospitalization and these infections may be life-threatening (1). Vaccines against pertussis first came to the market in the 1950s. The Netherlands has included pertussis vaccination in its National Immunization programme (NIP) since 1954, and this led to a sharp decline in pertussis fatalities (2, 3). Over seven decades several changes have been made to the vaccination schedule as vaccine technology advanced. Most notable has been the switch from whole cell vaccines to acellular vaccines, which was implemented in the Netherlands in 2005 (2, 4). To prevent severe disease in newborns, maternal vaccination at 22 weeks gestational age has been introduced in 2019 (2, 4). The current schedule thus consists of maternal vaccination, followed by a 2+1 scheme at 3 months, 5 months and 12 months, and a booster at 5 years of age. In case of insufficient protection from maternal vaccination the infant receives a 3+1 scheme at 2 months, 3 months, 5 months and 12 months, and a booster at 5 years of age (2, 4). The primary series currently uses a five component vaccine containing pertussis toxoid (PT), filamentous hemagglutinin (FHA), pertactin (PRN) and both fimbriae (fim2 and fim3), while the booster dose is a three component vaccine that lacks the fimbriae.

Many countries have switched from whole cell vaccines to acellular vaccines from the 1990s onwards, and this has led to vaccine-driven evolution of the bacterium(5, 6). The most notable has been an increase in pertactin-deficient strains in many countries (7). Upon the switch to acellular pertussis vaccines the Netherlands began to monitor trends in vaccine antigen expression amongst Dutch *Bordetella pertussis* isolates. Diagnostic laboratories around the country are encouraged to send any isolates they cultured or any clinical specimen found positive upon molecular testing to the reference laboratory for the pathogen surveillance programme.

During the COVID-19 pandemic and associated non-pharmaceutical interventions, diagnoses of whooping cough were at an all-time low (8, 9). Dutch epidemiological data (2) and a cross-sectional study primarily targeted to detection of SARS-CoV-2 [unpublished data] have both indicated a lack of circulating *Bordetella pertussis* prior to 2023. Since 2023 most European countries have reported a re-emergence of pertussis. This was accompanied by record high incidence rates and significant numbers of fatalities, as well as the appearance of macrolide resistant pertussis strains in Europe (10-14).

The Dutch pertussis reference laboratory started receiving samples in the fall of 2023 and this continued throughout 2024; >1000 samples were received for which culture was attempted. Culturing *Bordetella spp*. is notoriously insensitive, nevertheless a total of 193 pure cultures were obtained and typed for surveillance purposes. Here, we describe the Dutch *Bordetella pertussis* isolates of the 2023-2024 epidemic. We focus predominantly on expression of vaccine antigens, and the genetic variation in the corresponding genes. Using cgMLST 180/193 recent Dutch isolates were placed in context of strains circulating prior to the COVID-19 pandemic, as well as isoaltes from the past year from other European countries.

## Materials and methods

### Culture

Upon a positive molecular diagnostic test, diagnostic laboratories either inoculate Bordetella Selective medium plates (ThermoFisher) and sent these to the reference laboratory, or directly sent positive swabs – after which plates are inoculated at the reference laboratory. Plates are incubated at 35°C and high humidity for a maximum of 14 days. Presence of presumptive *Bordetella* colonies is checked every other day, species of two colonies is confirmed by Maldi-ToF (Bruker) and a single colony culture is started on a Bordet Gengou plate (BD BBL). Bordet Gengou plates are incubated for a maximum of seven additional days at 35°C and high humidity. Isolates are stored at −70°C and in parallel processed for whole genome sequencing.

### Whole genome sequencing and bio-informatics

Bacterial DNA was purified using the DNeasy blood and tissue kit (Qiagen) according to the manufacturer’s protocol “Purification of Total DNA from Animal Tissues (SpinColumn Protocol)” including a pretreatment for Gram-negative bacteria, or with the Maxwell RSC Cultured Cells DNA Cartridges (Promega) according to the manufacturer’s protocol. The Illumina DNA Prep kit (Illumina, Inc.) was used for library preparation.

Sequencing was performed on an Illumina NextSeq 550 using a NextSeq 500/550 Mid Output Kit v2.5 (150 Cycles) (Illumina, Inc.) or the NextSeq 2000 using NextSeq 1000/2000 P1 Reagents, both resulting in 2 × 150 bp reads. Read quality control, *de novo* assembly and genome assembly quality control were integrated in the juno-assembly pipeline v2.2.4 (15), which uses Spades (16) in isolate mode. Multilocus sequence typing (MLST) and Bordetella vaccine antigen typing are part of the juno-typing pipeline v0.8.6 (17). The integrity of the PRN gene was checked in the CLC Genomics Workbench (Qiagen), using the whole-genome alignment module and the PRN gene of the B1917 strain and it’s promoter region as a reference.

### WGS data analysis (BIGS-DB)

Genome assemblies of 2023-2024 isolates were selected for upload on the BIGS-DB Pasteur platform for further analysis (BIGS-DB ids 6220-6399). Duplicate isolates from the same (anonymized) patient were excluded from the dataset, as were genome assemblies not meeting BIGS-DB quality criteria; contigs > 500 and genome size 3.300.000 – 4.100.000 (n=13/193). Vaccine antigen typing, type III secretion system typing and core genome MLST (cgMLST) typing were performed automatically by BIGS-DB upon upload of the genome assemblies. The genome comparator module (18) was used to determine allelic distances between isolates. The minimum spanning tree was generated and visualized using the GrapeTree plug-in on the BIGS-DB platform (19).

### Statistics

Provenance and typing data of all isolates received between 2015-2024 was analysed using R (v4.4.1) and tidyverse (v2.0.0). Odds ratios (OR) were calculated using the epi.2by2 function of the epiR package (v2.0.68). OR were considered statistically significant when the 95% confidence interval (95%CI) excluded 1, this corresponded with Fisher’s exact test p-values of at least *p*≤0.03. Haldane-Anscombe correction was used to approximate the OR when one of the cells in a 2-by-2 table had 0 observations.

## Results

### Characteristics of 2023-2024 pertussis isolates

Consistent with trends in many other European countries (7, 20-22), the Netherlands observed a modest increase in pertactin deficient isolates in the period of 2015-2020. Thirteen percent of isolates was pertactin deficient in 2015-2017, this increased to 25% in 2018-2020 (OR 2.22; 95% CI 1.19 – 4.13). In contrast, 93% of isolates in 2023-2024 expressed pertactin (OR 0.29; 95% CI 0.15 – 0.57, Fig 1A). Dutch pertussis isolates from 2023-2024 belonged to one of three main genotypes; 54% ptxP3/fim3-1, 28% ptxP3/fim3-2, 13% ptxP1/fim3-26 (Fig 1B). Isolates from previous years (2015-2020) belonged predominantly to either ptxP3/fim3-1 or ptxP3/fim3-2. Prior to the COVID-19 pandemic, isolates showed an increase of the ptxP3/fim3-2 genotype over time (54% in 2015-2017 vs 67% in 2018-2020; OR 1.74; 95% CI 1.07 – 2.82). One 2024 isolate had an A→T mutation in the start codon of fim3 but was otherwise identical to fim3-2, this variant has been designated allele 49 in BIGS-DB Pasteur.

**Figure 1:**
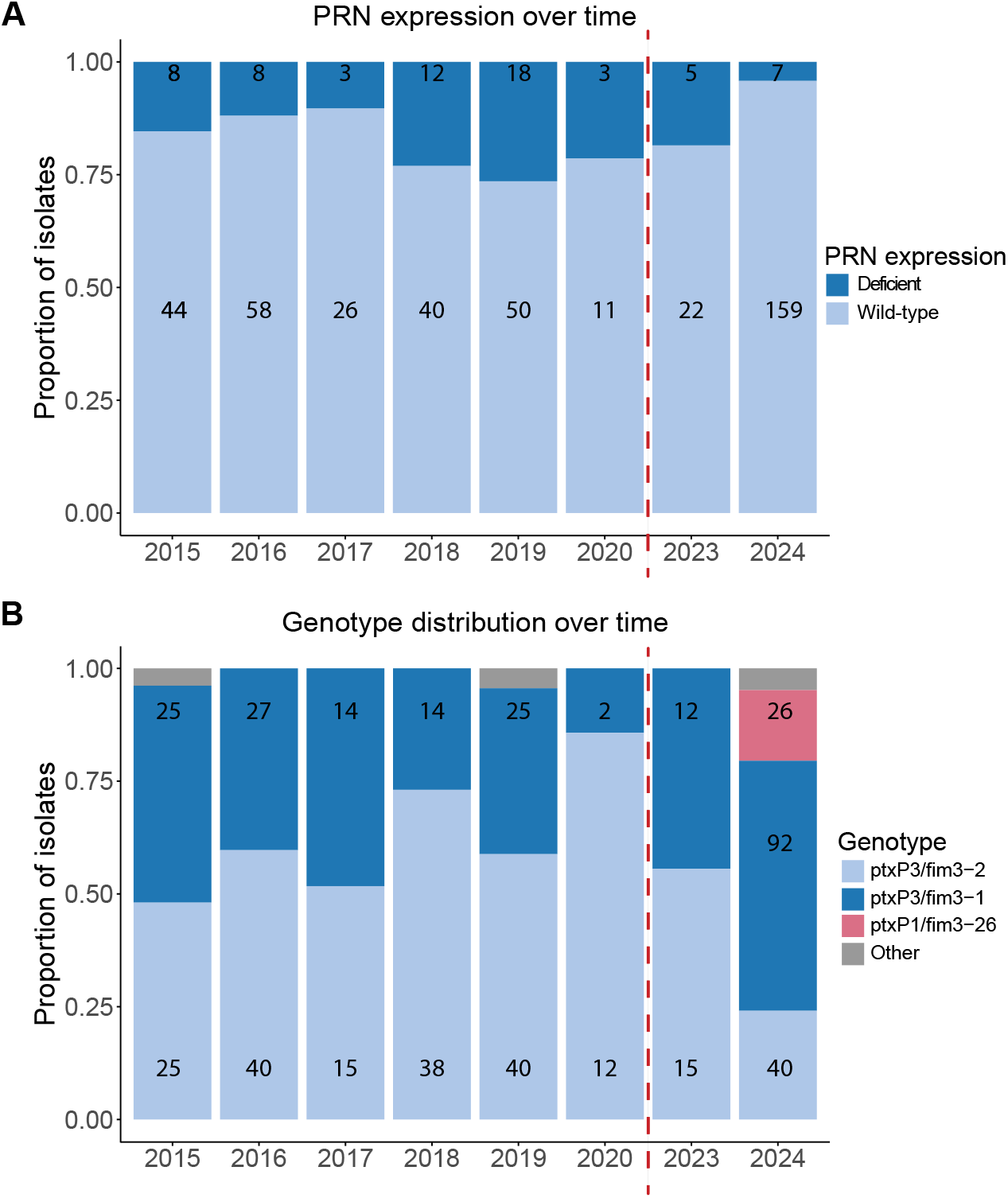
Pertactin expression and genotype distribution over time. Proportion and absolute number of pertussis isolates with pertactin deficiency **(A)** and per genotype **(B)** over a 10 year time period, plotted per year. Absolute number of isolates written in the bars. Dashed red line indicates the COVID-19 pandemic period, when no samples were received by the reference laboratory.

Amongst isolates from 2015-2020 of genotype ptxP3/fim3-1 30.4% was identified as pertactin deficient. This was significantly less common amongst isolates of the ptxP3/fim3-2 genotype, where only 9.1% was pertactin deficient (OR 0.27; 95% CI 0.14 – 0.50). The isolates of the ptxP1/fim3-26 genotype were all isolated in 2023-2024, all isolates of this genotype expressed pertactin. Thus far, no Dutch pertussis isolates carry the A2047G mutation in the 23S rRNA gene conferring macrolide resistance.

### Novel allele of T3SS translocator gene *bopD* found, strain in circulation since fall of 2023

Twenty-one Dutch isolates of genotype ptxP3/fim3-2 carry a C151T mutation in the *bopD* gene, resulting in a P51S substitution in the protein. *bopD* encodes the translocator protein of the type III secretion system (T3SS), and as such is part of an established bacterial virulence system. The effect of the C151T mutation on protein functionality remains unclear at this moment. The twenty-one isolates have been received between October 2023 and July 2024, from different geographical locations and from both children and adults; all indicative of stable circulation of this strain throughout the Netherlands during the pertussis epidemic. The *bopD* variant was not detected amongst 135 pertussis isolates from 2018-2020 (OR 36.53; 95% CI 2.19 – 608.69).

### Novel genotypes show less genetic diversity compared to stably circulating genotypes

Genotypes carrying ptxP1 are considered ancestral, while the ptxP3 allele has dominated the pertussis population in the acellular vaccine era. To explain the emergence of the ptxP1/fim3-26 genotype in the past epidemic we investigated the genomic diversity using a previously described cgMLST approach of 2038 loci (23). At this higher resolution 178/180 of the Dutch pertussis isolates of 2023-2024 had a unique allelic profile. The remaining two isolates were typed as the same strain; these were both isolated in February of 2024 but with significant geographical spread (100km apart). One was from a one-year-old patient and the other from a 16-year-old patient.

The mean overall genetic diversity over all 180 genome assemblies was 38.5 allelic differences (AD). Within a genotype the diversity was, by definition, lower. This was found to be 25.6 AD (ptxP3/fim3-1 (VA-ST4), n=99), 29.3 AD (ptxP3/fim3-2 (VA-ST9), n=54), and 18 AD (ptxP1/fim3-26 (VA-ST93), n=23) respectively.

In a minimum spanning tree representation of the cgMLST data (Fig 2) the different levels of genetic diversity within the classical genotypes can be observed. The genotypes with a higher internal diversity ptxP3/fim3-1 (dark blue) and ptxP3/fim3-2 (light blue) split into multiple clusters, while the ptxP1/fim3-26 isolates (orange) appear as one cluster. The isolates with the *bopD* mutation (labelled with “*” in the spheres) are predominantly of the ptxP3/fim3-2 genotype and appear as one cluster within this genotype, suggestive of this being a lineage. The average genomic diversity amongst these bopD mutant isolates is 21 AD.

**Figure 2:**
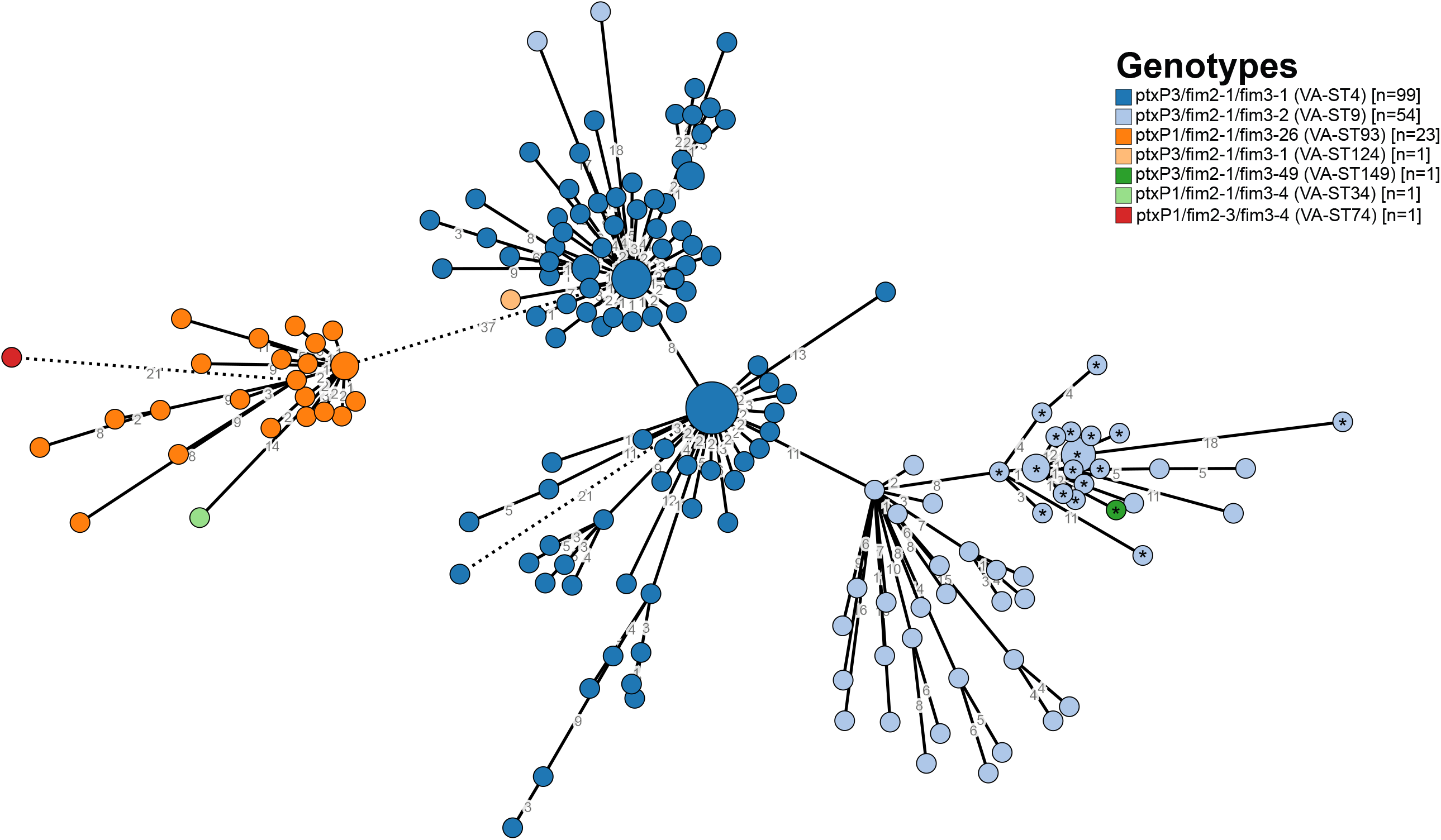
Minimum spanning tree based on cgMLST typing. Pairwise distances excluding missing alleles were used to determine a minimum spanning tree between unique cgMLST profiles. The numbers on the branches indicate the number of allelic differences between cgMLST profiles and branches longer than 11 AD have been drawn as dotted lines. Each circle represents a cgMLST profile (excluding missing loci), the diameter of circles represents the number of isolates of this profile. Colours represent the different genotypes; ptxP1/fim3-26 in orange, ptxP3/fim3-1 in dark blue and ptxP3/fim3-2 in light blue. Isolates with the novel *bopD* allele are label with *. cgMLST: core genome multilocus sequence typing; AD: allelic distance

## Discussion

Many European countries observed a resurgence of whooping cough in 2023 and 2024. Typing results of Dutch pertussis isolates from this resurgence were largely consistent with those reported by other European countries (11, 13); almost all isolates express pertactin and strains of the ptxP1 genotype are circulating, resulting in overall increased genetic diversity in the Dutch pertussis population.

Pertussis with a ptxP1 promoter was shown to have reduced fitness in the acellular vaccine (ACV) era, in comparison to ptxP3 strains (24). This made the circulation of such strains in several European countries unexpected and noteworthy. During and after the COVID-19 pandemic circulation of *B. pertussis* has been extremely low for several years (2, 8, 9). The lack of circulation – and thus boosting of vaccine-induced immunity after exposure – may have partly shifted the fitness balance. Pathogen surveillance data of upcoming pertussis seasons will show whether this trend continues or whether European pertussis isolates will shift back to be dominated by ptxP3 genotypes. The ptxP1 strain circulating in the Netherlands (ptxP1/fim3-26) showed moderate genetic diversity with a mean allelic distance of 18. Five recent isolates of the same genotype (BPagST93) were available on the BIGS-DB platform hosted at Institut Pasteur, these were isolated in France (n=4) and Belgium (n=1). The average allelic distance of Dutch isolates to these international isolates was 26 AD. The minimum difference between the Belgium isolate and a Dutch isolates was only 1 AD, while the closest French isolate was 10AD apart. This international context suggests that the genetic diversity amongst the ptxP1/fim3-26 genotype is quite limited. This, combined with the modest detection of the genotype in other countries, may suggest a single introduction of the ptxP1/fim3-26 genotype to the European pertussis population. Whether this was a travel-induced re-introduction from elsewhere or the unmasking of an old European ptxP1 strain remains to be investigated.

Several European countries have reported macrolide resistant *Bordetella pertussis* (MRBP) isolates (11, 13, 25). MRBP was first described in 1994 (26) but has become much more common since ∼2010 (27-31). The most commonly found resistance mechanism is an A2047G mutation in the 23S rRNA gene (27, 28, 32-35). MRBP isolates were initially of the ptxP1 genotype (29), but a more recent MRBP strain was found to be of the ptxP3 genotype (33-35), including those MRBP isolates found in Europe (11, 13). Thus far none of the Dutch pertussis isolates have harboured the 23S rRNA mutation.

Instead, we report a novel variant of the *bopD* gene that was found in 12% (21/180) of the genome assemblies of recent Dutch pertussis isolates. This variant of *bopD* was not found in any other isolate when searching the Institut Pasteur BIGS-DB database, which contained over 3000 public genomes at the time of writing, thereby suggesting this variant to be a unique Dutch *B. pertussis* lineage. The Bordetella T3SS injects BteA (also known as BopC) into the cytoplasm of the host cell where it triggers cell death (36-38). The regulator BopN also ends up in the host cell cytoplasm, it was shown to interfere with NF-κB signalling and upregulate the anti-inflammatory cytokine IL-10 (36, 39, 40). BopD is one of the translocators of the T3SS, together with BopB (36, 41). An alanine insertion into BteA of *B. pertussis* was shown to reduce the cytotoxicity when compared to *B. bronchiseptica* BteA (36, 42). The P51S substitution in BopD is located in the N-terminal part of the protein (gene length 942). Unfortunately there is no structural information on BopD and thus it is hard to predict the effect of the substitution on protein functionality. As the 21 Dutch isolates carrying this mutation have been isolated over an eight-month time period and from geographically diverse locations, the mutation is unlikely to hamper virulence or bacterial spread. Surveillance data from other countries may provide further information on the spread of pertussis carrying this novel allele. Meanwhile, molecular biological work may elucidate the effect of the point mutation on protein functionality.

Continued surveillance and monitoring of *B. pertussis* isolates remains essential; to monitor trend in serotypes and genotypes, to detect new variants of virulence factor genes, and for the timely detection of MRBP isolates. Primary pertussis diagnostics in the Netherlands is culture-independent and instead relies on serology or molecular detection of the bacterium. As such, detection of newly arising variants, albeit variants with changes in virulence factors or MRBP, relies on the national pathogen surveillance programme.

## Data Availability

Whole genome sequencing data with limited provenance data will be made available on the BIGS-DB Bordetella platform.

## Acknowledgments

We acknowledge the contribution of participating diagnostic laboratories in the Netherlands to the pathogen surveillance programme. We wish to thank Dimphey van Meijeren and Dr. Casper Jamin for critically reading the manuscript and scientific discussion.

